# Respiratory syncytial virus-specific immunoglobulin G (IgG) concentrations associate with environmental and genetic factors: the Factors Influencing Pediatric Asthma Study

**DOI:** 10.1101/2021.08.17.21262198

**Authors:** Esther Erdei, Dara Torgerson, Rae O’Leary, Melissa Spear, Matias Shedden, Marcia O’Leary, Kendra Enright, Lyle Best

**Affiliations:** University of New Mexico Health Sciences Center College of Pharmacy, Albuquerque, NM, USA; Department of Epidemiology and Biostatistics, University of California San Francisco, San Francisco, CA, USA; Department of Human Genetics, McGill University, Montreal, QC, Canada; Missouri Breaks Industries Research Inc, Eagle Butte, SD, USA; Turtle Mountain Community College, Belcourt, ND, USA

## Abstract

Exposure to respiratory syncytial virus (RSV) during childhood is nearly ubiquitous by age two, and infants who develop severe RSV bronchiolitis are more likely to develop asthma later in life. In the Factors Influencing Pediatric Asthma (FIPA) study including 319 children from a Northern Plains American Indian community, we found 73% of children to have high concentrations of RSV-specific IgG (>40 IU/mL). High concentration of RSV-specific IgG was associated with increased exposure to second-hand tobacco smoke (p=7.5×10^−4^), larger household size (p=4.0×10^−3^), and lower levels of total serum IgE (p=5.1×10^−3^). Parents of children with asthma more often reported an RSV diagnosis and/or hospitalization due to RSV, and children with asthma had lower concentrations of RSV IgG as compared to those without asthma among RSV-exposed individuals (mean 117 IU/mL vs. 154, p=7.1×10^−4^). However, lower RSV IgG was surprisingly exclusive to children with asthma recruited during the winter months when RSV is thought to circulate more broadly. Multivariate regression indicated the strongest predictors of RSV-specific IgG concentration included asthma status (p=0.040), per cent eosinophils (p=0.035), and an asthma x RSV season interaction (p=3.7×10^−3^). Among candidate genes, we identified a genetic association between an intronic variant in *IFNL4* and RSV-specific IgG concentration whereby the minor allele (A) was associated with higher concentration (rs12979860, p=4.3×10^−3^). Overall our findings suggest there are seasonal differences in immunological response to RSV infection in asthma cases vs. controls, and identify both environmental and genetic contributions that warrant further investigation.

## Introduction

Respiratory syncytial virus (RSV) is an RNA virus, classified as a member of the Paramyxoviridae family of negative-strand RNA viruses in the order Mononegavirales. It is the most common cause of bronchiolitis in infants during the first 2-3 years of life [1]. Infection by the human RSV orthopneumovirus is ubiquitous, affecting nearly all children by two years of age worldwide [1]. The incidence of hospitalization for bronchiolitis in European and American populations is about 30 per 1,000 in the first year of life [2,3]. Ethnic differences in incidence are noted in the United States, with increased risk among Hispanic and American Indian populations [4,5]. Severe RSV infection as an infant confers increased susceptibility to asthma [6–8] and perhaps atopy [6], within at least the subsequent two decades of life. There is also evidence of impaired lung function among young adults with viral bronchiolitis as infants [9]. Almost all infectious agents are involved in complex interactions within their host, and this seems especially true for RSV infection and sequelae. There is evidence that host genetic factors influence the initial pathogenesis of RSV infection [10–13], and that the development of asthma following severe RSV infection is likely due to both genetic [11] and environmental [14] contributions. However, the relationship between immunological response to RSV infection and asthma remains largely unexplored.

In spite of pervasive childhood infection and persisting IgG antibodies to RSV in adulthood [15,16], recurrent RSV infection among adults is relatively common [17–19] and a cause of significant morbidity [20,21]. In Tribal community settings, where multigenerational households are common, understanding RSV infection burden from infancy to adulthood is especially important to design preventions and protect community health. The Factors Influencing Pediatric Asthma (FIPA) study [22] enrolled 324 children from a Northern Plains American Indian community to characterize the environmental, social and genetic factors associated with asthma in this high-risk, high-prevalence community. The goal of the present analysis was to quantify the immunoglobulin G response to RSV infection in American Indian children with and without asthma, and to investigate the environmental, clinical, and genetic factors that associate with varying response.

## Methods

### Ethics approval and consent to participate

The research protocol was approved by the institutional review boards at the Collaborative Research Center for American Indian Health, Sanford Research, Sioux Falls, SD; the Great Plains Indian Health Service IRB, Aberdeen, SD and the local Tribal government. All participants’ parents gave informed consent in writing and all children provided assent.

### Study population

The Factors Influencing Pediatric Asthma (FIPA) study, enrolled 324 participants from a single site in a population-based, case/control study as previously described [22], of which 319 were included in the current study. Briefly, 108 asthma cases were ascertained by a global search of Indian Health Service electronic medical records for a diagnosis related to asthma (ICD codes 493.00 through 493.92, plus 786.07 [wheezing] and V17.5 [family history of asthma]), and more specifically defined by at least 2 of 3 potential criteria (asthma diagnosis, at least 2 prescriptions for bronchodilator medications within the past 2 years, or pulmonary function testing showing a 20% improvement in FEV1 post bronchodilator treatment). Participating children were between 6 and 17 years of age, and two asthma controls were recruited for every asthma case. Cases were individually age-matched (within 6 months, N=216) to control children who did not meet case criteria, primarily by a similar search of electronic medical records.

Of 319 participants, 15 indicated their child was born prior to 36 weeks gestational age including 6 asthma cases and 9 asthma controls (minimum reported gestational age of 32 weeks, ranging from 32 – 35 weeks). Palivizumab may in some cases be administered in the NICU to infants born prematurely to provide immunoprophylaxis against RSV, which may alter immunological response to RSV infection. However, we were unable to query NICU records to obtain this information, and parents were not specifically asked whether their child had ever received Palivizumab in infancy.

### Study procedures

Parents of potential participants were contacted by study field assistants via telephone and invited to participate if eligibility criteria were met. Parents of all participants completed a standardized questionnaire related to socio-demographic, housing environment and health history of their children. Two questions related to prior RSV infections were asked, including “Has a medical person ever said that your child had RSV? Yes or No.”, and “Has your child ever had to stay overnight in the hospital? Yes or No.”. If they responded yes to prior hospitalization, parents were asked to fill in the “Reason/Diagnosis”, and “Age”. From these questions, we generated two variables related to clinical manifestation of RSV infection, including self-reported “RSV infection” and “RSV hospitalization”.

At baseline, all children were examined for anthropometric measures, pulmonary function testing, and home environmental sampling. The following laboratory samples were obtained: (1) Blood was collected to perform basic measures of immune status including % eosinophils, RSV IgG, total serum IgE, and C-reactive protein (CRP), 2) salivary DNA samples (Oragene OGR-500, DNA Genotek, Ottawa, Canada) were collected for DNA genotyping, following manufacturer approved extraction, and (3) salivary cotinine samples (Accutest, NicAlert, Encino, California) were collected to measure exposure to tobacco products in the past 48 hours. A follow-up exam approximately 3 years later, repeated pulmonary function testing and collected additional biological samples for ancillary testing as described above.

### Laboratory measurements

**RSV-specific IgG serum antibody concentrations** were determined on 319 participants, using stored serum collected at either baseline (1^st^ exam, or study entry, N=48) or at follow-up (2^nd^ exam, N=271). RSV IgG serum concentration was quantified using an IBL International certified sandwich ELISA assay (Hamburg, Germany, catalogue number: RE RE56881-10, Lot#: RG-204). Each serum sample was diluted 1:100 times before use, and 100 µL of each Standard solution (A-D) and diluted sample were added to the RSV antigen pre-coated wells of the microtiter plate. The process followed the protocol provided by the company and quality control standards were included and assured at each run. Optical density (OD) was measured with a spectrophotometer at 450 nm (Reference-wavelength: 600-650 nm) within 60 min after pipetting of the stop solution. Standard curve point-to-point analysis method was applied for each plate and run to determine RSV-specific IgG concentrations (IU/ml) for each participant’s serum sample.

**Measurement of high sensitivity C-reactive protein** (CRP) in serum was done at Sanford Laboratories, Rapid City, SD, using a particle enhanced immunoturbidimetric assay (Roche dedicated CRPHS reagent, catalog number 04628918190, Roche Diagnostics, Indianapolis, IN), on a Roche Cobas 6000 instrument using the manufacturer’s recommended protocols.

**Total IgE_**determinations were also carried out by Sanford Laboratories, (Sioux Falls, SD) using fluorescent enzyme immunoassays for each antigen, (ImmunoCAP dedicated reagents, catalog number 10-9319-01, Phadia AB, Uppsala, Sweden) on the Phadia ImmunoCAP 250 instrument.

**Complete blood count**, including eosinophil and other specific cells, was measured on a SYSMEX XNL-450 hematology analyzer (Lincolnshire, IL) at the local hospital within hours of phlebotomy, using reagents and quality controls as specified by the manufacturer.

**Salivary cotinine** measures were obtained using the Accutest NicAlert™ (Jant Pharmacal Corp, Encino, CA) with visual comparison to a standard strip resulting in 0-7 ordinal levels.

**Genome-wide SNP genotyping** was performed for 2.5 million variants on the expanded multi-ethnic genotyping array (MEGA^EX^ array, Illumina Inc.), with standard variant calling and quality control measures applied (genotyping rate > 95%, Hardy-Weinberg p>0.0001, tests for unexpected relatedness/duplication between samples). The present study examined 11 selected variants related to clinical RSV outcomes. These additional measures were consistent with the participants’ consents and approved by Tribal authorities.

### Statistical analysis

Statistical analyses were performed using R [23]. Contrasts of the clinical characteristics of asthma cases vs. controls, study participants with high IgG (>40 IU/ml) vs. low/undetected IgG (<40 IU/ml), and individuals with asthma reporting asthma medication usage were performed using a student’s t-test. Multi-variable linear regression was used to evaluate the correlation between serum RSV-specific IgG concentrations (IU/ml) and asthma, household size at study entry, body mass index (BMI), gender, acute exposure to tobacco (using saliva cotinine determinations), serum concentration of CRP, age, total IgE, and % eosinophils in whole blood.

The effect of seasonality on RSV-specific IgG concentration was also evaluated by generating a binary variable for serum samples collected during periods of potentially higher RSV transmission (December – April) vs. lower RSV transmission (May – November), at either the baseline or follow-up visit. Tests of genetic association with 11 candidate variants selected from the literature potentially associated with RSV immune or clinical response were performed using linear regression, adjusting for asthma, age, and sex. Values of RSV-specific IgG concentration that were below assay detection (left-censored) were transformed using 1√2 (N=11 samples) prior to association testing.

## Results

Quantitative measurements of serum RSV IgG were obtained among 108 individuals with asthma, and 211 individuals without asthma (total N = 319). Clinical and demographic characteristics of asthma cases and controls are summarized in Table 1. At study entry, mean age, % male gender, and levels of C-reactive protein (CRP) were not significantly different between asthma cases and controls. However, individuals with asthma had a significantly smaller household size at baseline as compared to asthma controls. (Table 1; p=1.6×10^−4^). Asthma cases also had higher levels of total serum IgE as compared to asthma controls (Table 1, p=3.1×10^−5^), and a non-significant trend towards having higher BMI (p=0.054). The geometric mean of total IgE was 111 and 53% of children were above the clinical upper limit of normal (100 kU/L).

**Table 1.** Clinical characteristics of asthma cases and controls with measured serum RSV IgG in the Factors Influencing Pediatric Asthma study (FIPA).

|  | **Asthma Cases** | **Asthma Controls** | **P-value** |
| --- | --- | --- | --- |
| **N** | 108 | 211 | - |
| **Age at Enrollment (mean)** | 11.8 | 12.2 | 0.32 |
| **Gender (% male)** | 52.8 | 50.7 | 0.81 |
| **BMI (mean)** | 25.4 | 23.7 | 0.054 |
| **Cotinine (NicAlert, mean)** | 0.99 | 1.17 | 0.098 |
| **Household Size at Enrollment (mean)** | 5.3 | 6.3 | $1.6 \times 10^{-4a}$ |
| **Undetected (IgG < 10 IU/mL, N)** | 0 | 26 | $1.6 \times 10^{-5a}$ |
| **High IgG (IgG > 40 IU/mL, %)** | 61.1 | 80.6 | $2.6 \times 10^{-4a}$ |
| **RSV IgG (IU/mL, mean)** | 117 | 135 | 0.093 |
| **RSV IgG (IU/mL, mean, excluding undetected)** | 117 | 154 | $7.1 \times 10^{-4a}$ |
| **Hospitalizations from RSV (%)** | 17.6% | 6.7% | 0.00358<sup>a</sup> |
| **RSV diagnosis (%)** | 34.6% | 16.1% | $4.5 \times 10^{-4a}$ |
| **Total Serum IgE [log (kU/L), mean]** | 5.2 | 4.4 | $3.1 \times 10^{-5a}$ |
| **% Eosinophils (mean)** | 5.0 | 3.8 | 0.0098 |
| **CRP (log, mean)** | 0.16 | 0.0050 | 0.22 |

<sup>a</sup>Statistically significant following Bonferroni adjustment for 14 tests ( $p < 3.66 \times 10^{-3}$ ).

The majority of individuals were classified as having high RSV-specific IgG concentrations (N=236, or 73%), which was defined as being above 40 IU/ml threshold by validation against the WHO standard per company protocol. A total of 26 individuals had RSV-specific IgG concentrations < 10 IU/ml, indicating either no prior infection/exposure to RSV or an impaired humoral response. All of these individuals were asthma controls. To confirm the quality of the 26 serum samples where no or low RSV IgG levels were detected, we measured vaccination-induced IgG concentrations against measles, pertussis, and tetanus antigens (IBL International detection kits used). All samples had measurable concentrations expressed as IU/ml values, providing additional evidence these samples were of high quality and reflect reliable measures of no/low RSV-specific IgG. Furthermore, we found vaccine-induced IgG concentrations were not significantly different in individuals with low/no RSV IgG as compared to 17 individuals with RSV IgG concentrations > 10 IU/ml (S1 Fig). Characteristics of individuals with high as compared to low concentration of RSV-specific IgG are presented in Table 2, which include higher levels of cotinine (p=8.6×10^−4^), larger household sizes at enrollment (p=4.0×10^−3^), and lower levels of total serum IgE (p=5.1×10^−3^).

**Table 2.** Comparison of individuals with high RSV IgG (>40 IU/ml vs. low (<40 IU/ml).

|  | High IgG | Low IgG | P-value |
| --- | --- | --- | --- |
| **N** | 236 | 83 | - |
| **Age at Enrollment (mean)** | 12.1 | 11.8 | 0.38 |
| **Gender (% male)** | 50.4 | 54.2 | 0.61 |
| **BMI (mean)** | 24.1 | 24.7 | 0.46 |
| **Cotinine (NicAlert, mean)** | 1.20 | 0.855 | $7.5 \times 10^{-4a}$ |
| **Household Size at Enrollment (mean)** | 6.2 | 5.4 | $4.0 \times 10^{-3a}$ |
| **Total Serum IgE [log (kU/L), mean]** | 4.57 | 5.12 | $5.1 \times 10^{-3a}$ |
| **CRP (log, mean)** | 0.33 | 0.27 | 0.65 |

<sup>a</sup>Statistically significant following Bonferroni correction for 7 tests ( $p < 7.1 \times 10^{-3}$ )

In our contrasts of individual variables, we found asthma cases had significantly lower serum RSV-specific IgG concentrations as compared to asthma controls among exposed individuals (p=7.1×10^−4^, Table 1). Parents/guardians of asthma cases were more likely to report their child had been previously diagnosed with RSV by a healthcare professional as compared to parents/guardians of asthma controls (34.6% vs. 16.1% respectively, p=4.5×10^−4^), more likely to report hospitalization due to RSV infection (17.6% for asthma cases vs. 6.7% for asthma controls, p=0.0036).

Asthma cases recruited during typical RSV season (December – April) had significantly lower total RSV-specific IgG as compared to asthma cases recruited outside of RSV season (May – November), and as compared to asthma controls regardless of RSV season (Fig 1). However, apart from RSV-specific IgG concentrations, we found no differences in the clinical and demographic variables between asthma cases recruited during vs. outside of RSV season (Table 3). In contrast, asthma controls had similar levels of RSV IgG irrespective of the season they were recruited in. In our multi-variate regression model, variables that were associated with RSV-specific IgG were limited to asthma, % of blood eosinophils, and the interaction between asthma x RSV season (Table 4).

**Figure 1.**
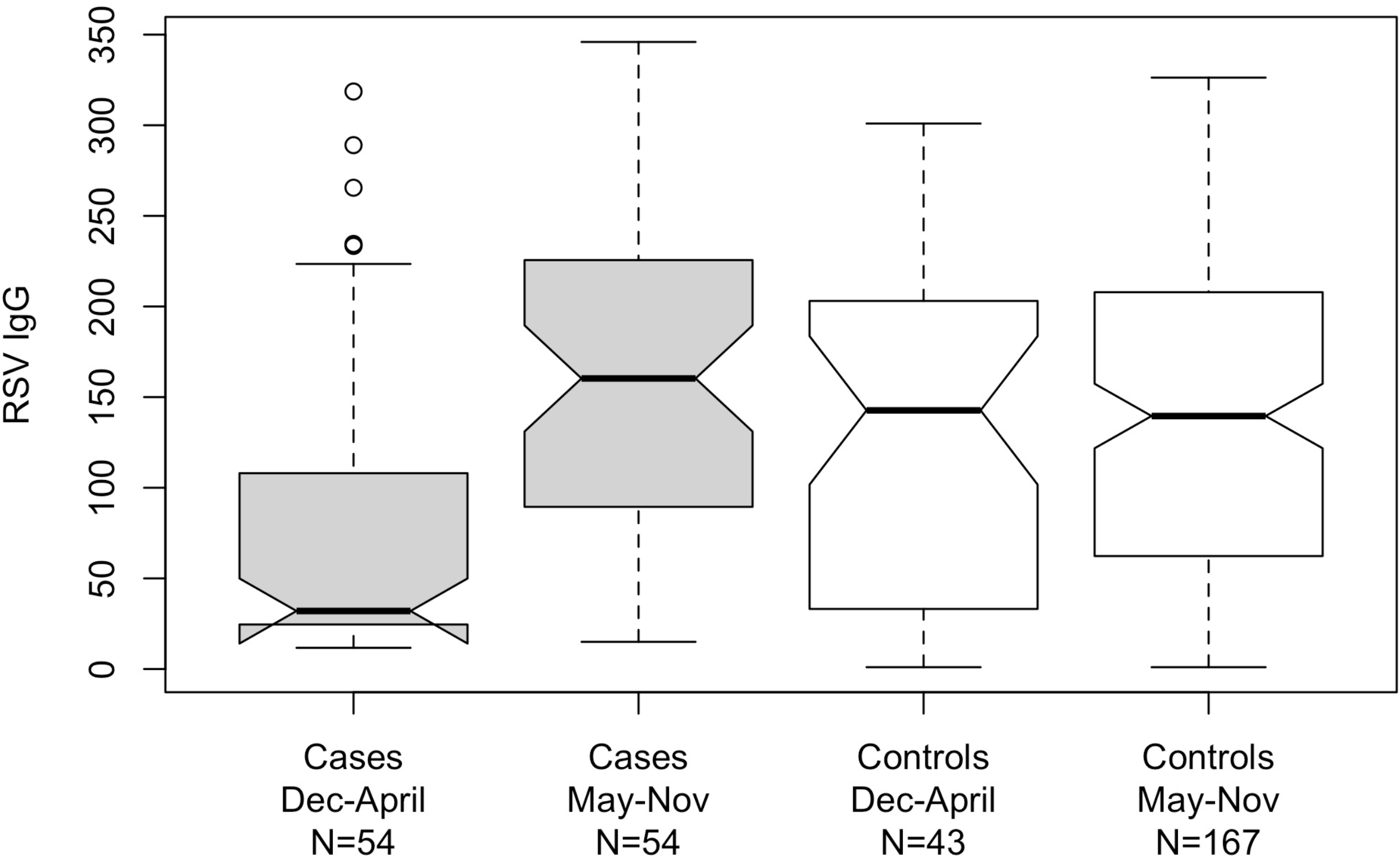
Boxplot showing the distribution of serum RSV IgG in asthma cases vs. controls, stratified by RSV season. Asthma cases recruited during RSV season (Dec – April) have significantly lower RSV IgG as compared to asthma cases recruited outside RSV season (May – Nov) (t-test, p=2.5×10^-6^). There was no observed difference in seasonality for asthma controls (p=0.60).

**Table 3.** Clinical characteristics of asthma cases recruited during the winter/spring (December through April) when RSV is typically circulating more broadly in the community vs. the summer/fall (May through November).

|  | Winter/Spring | Summer/Fall | P-value |
| --- | --- | --- | --- |
| **N** | 54 | 54 | - |
| **Age at Enrollment (mean)** | 12.0 | 11.6 | 0.53 |
| **Gender (% male)** | 53.7 | 51.9 | 1.0 |
| **BMI (mean)** | 26.0 | 24.9 | 0.47 |
| **Cotinine (NicAlert, mean)** | 0.85 | 1.1 | 0.13 |
| **Household Size at Enrollment (mean)** | 5.0 | 5.6 | 0.13 |
| **Undetected (IgG<10 IU/mL, N)** | 0 | 0 | 1.0 |
| **High IgG (IgG > 40 IU/mL, %)** | 37.0 | 85.2 | 4.3x10<sup>-7a</sup> |
| **RSV IgG (IU/mL, mean)** | 75.8 | 157 | 2.5x10<sup>-6a</sup> |
| **Hospitalizations from RSV (%)** | 18.5% | 16.7% | 1.0 |
| **RSV diagnosis (%)** | 33.3% | 35.8% | 0.84 |
| **Total Serum IgE [log (kU/L), mean]** | 5.3 | 5.1 | 0.53 |
| **% Eosinophils (mean)** | 4.9 | 5.1 | 0.80 |
| **CRP (log, mean)** | 0.038 | 0.38 | 0.26 |

<sup>a</sup>Statistically significant following Bonferroni adjustment for 14 comparisons ( $p < 3.9 \times 10^{-3}$ ).

**Table 4.** Results of multiple linear regression for total serum RSV IgG concentrations.

|  | Estimate | Standard Error | Test Statistic | P-value |
| --- | --- | --- | --- | --- |
| **Asthma** | 29.8 | 14.4 | 2.07 | 0.0398<sup>a</sup> |
| **Household size** | 3.32 | 2.45 | 1.36 | 0.176 |
| **lnBMI** | -2.49 | 24.5 | -0.102 | 0.919 |
| **Gender** | -4.13 | 10.2 | -0.407 | 0.684 |
| **Cotinine (NicAlert)** | 6.76 | 5.90 | 1.15 | 0.253 |
| **lnCRP** | -4.44 | 5.76 | -0.770 | 0.442 |
| **age** | -1.83 | 1.90 | -0.959 | 0.339 |
| **RSV season** | -9.47 | 15.5 | -0.610 | 0.542 |
| **Total IgE** | 0.00108 | 0.00964 | 0.112 | 0.911 |
| **% Eosinophils** | -3.42 | 1.61 | -2.12 | 0.0350<sup>a</sup> |
| **Asthma x RSV season (interaction)** | -68.0 | 23.2 | -2.92 | 0.00373<sup>b</sup> |

<sup>a</sup> $p < 0.05$
<sup>b</sup> $p < 0.01$

Among 11 candidate variants selected from the literature to have potential effects on RSV infection, we identified a significant genetic association with an intronic variant in *IFNL4* (Interferon Lambda 4, Table 5).

**Table 5.** Results of genetic association at candidate variants with total serum RSV IgG concentration in 276 unrelated children with and without asthma. Tests for association were adjusted for asthma, age, and sex.

| Gene | SNPs | Allele (A1/A2) | Freq A1 | Beta A1 | Test Statistic | P-value | Clinical Impact (references) |
| --- | --- | --- | --- | --- | --- | --- | --- |
| *IFNG* | rs2430561 | T/A | 0.14 | -10.02 | -0.9861 | 0.3249 | RSV adult seroconversion [24,25] |
| *TNF* | rs1800629 | A/G | 0.038 | -9.539 | -0.4631 | 0.6437 | Severe influenza, Experi-RSV [25,26] |
| *IL-6* | rs1800795 | G/C | 0.098 | 7.457 | 0.6311 | 0.5285 | Experi-RSV, adults/children [24,25] |
| *VDR* | rs2228570 | G/A | 0.40 | -7.002 | -0.9072 | 0.3651 | RSV bronchiolitis, infants [27,28] |
| *MARCO* | rs1801275 | G/A | 0.36 | -3.211 | -0.4033 | 0.6871 | RSV and wheezing, Chilean children [29] |
| *IFNL4* | rs12979860 | A/G | 0.23 | 25.49 | 2.884 | 0.00425<sup>a</sup> | Lower age at hospital admission for bronchiolitis [30] |
|  | rs4986790 | G/A | 0.020 | 40.08 | 1.448 | 0.1487 | RSV in high risk infants [31]; severe RSV [10,32,33] |
| *IFNL3* | rs2243639 | A/G | 0.30 | 2.226 | 0.2791 | 0.7804 | RSV in Chilean infants [34] |
| *ADRB2* | rs1800888 | A/G | 0.0054 | 90 | 1.713 | 0.08782 | Asthma following severe RSV bronchiolitis[11] |
| *NOS1* | rs76090928 | A/G | 0.0018 | 92.8 | 1.022 | 0.3078 | Asthma following severe RSV bronchiolitis[11] |

<sup>a</sup>Statistically significant following Bonferroni adjustment for 11 statistical comparisons.

## Discussion

RSV and other viral infections at a young age have previously been associated with the later development of asthma [8,35–38]. In a study of American Indian children living in the Great Plain geographic area of rural South Dakota, we observed many similarities in the clinical characteristics of children with asthma as with prior studies, including increased BMI [39,40], increased hospitalizations or a diagnosis of RSV [41], higher levels of total serum IgE [42,43], and higher % eosinophils [44,45].

In the current study, we measured immunological response to prior RSV infection, and find children with asthma to have lower levels of serum RSV-specific IgG as compared to those without asthma. Surprisingly, our finding is specific to children with asthma who were recruited during the winter months, when RSV is thought to be circulating more broadly in the community and within households. Our findings suggest that children with asthma have a reduced or varying immunological response to RSV infection during the winter months as compared to children without asthma, which is not present during the summer months. Future longitudinal and prospective studies are warranted to determine the timing of exposure as it relates to IgG immunological response in children with and without asthma, and to determine any relationship with acute vs. long-term clinical outcomes.

Very few studies have examined RSV-specific IgG antibody levels in the context of asthma and/or bronchial hyperreactivity for comparison to our current results. Backer et al examined RSV-specific IgG among a randomly selected group of children between 7 and 16 years of age, but found no association with bronchial hyper-responsiveness to histamine challenge [46]. In another study among 100 children less than 2 years of age hospitalized for wheezing and followed for a median of 6 years, RSV-specific IgG increased with age, but was not associated with asthma at any age [47]. In our cross-sectional study, we found no significant association between RSV IgG concentration and age in either our single or multi-variate analysis, however we did not perform any longitudinal measurements within an individual. Although we are unaware of any recent findings to date, the INSPIRE longitudinal study of RSV serology among nearly 2,000 infants may provide further information as to whether immunological response to RSV infection is predictive of asthma or varies by season or age [48].

Lower RSV-specific IgG concentrations in asthma cases during the winter months when RSV is thought to be circulating more broadly in the community was an unanticipated finding (Fig 1). IgG antibodies typically peak within 3-4 weeks following RSV infection and gradually decline over the next few months [49,50]. Thus, we expected RSV IgG concentrations to be higher during times of recurring exposure during the winter months for both asthma cases and controls. While the seasonality of RSV infection is well accepted [50,51], we were unable to locate any independent cohort surveillance information or cross-sectional serology surveys that recorded seasonal timing of collection for comparison to our results. Thus, further efforts to validate our findings and identify potential causal factors driving the observed interaction between asthma and RSV season on IgG concentrations requires additional study.

Parents of children with asthma were more likely to report their child had been previously diagnosed with RSV, and we observed a non-significant trend towards increased rate of hospitalization due to RSV. While we do not have information on the clinical course of RSV infection in our study participants beyond self-report, we identified a shared genetic association that may provide additional insight as to whether RSV-specific IgG concentrations are of any clinical relevance. The *IFNL4* variant we found associated with RSV-specific IgG is a member of the interferon lambda family, which is known to be involved in the immune response to viral infection. We identified this as a candidate variant from Scagnolari et al [30], who found that individuals with the AA genotype had a lower age at hospital admission due to RSV infection as compared to infants carrying the GG/GA genotypes. In our study, Individuals with the AA genotype had significantly higher levels of RSV-specific IgG as compared to individuals with the GG genotype (Fig 2). Although this requires validation, our findings in combination with Scagnolari et al. [30] suggest that individuals with two copies of the minor allele (AA genotype) have a heightened immunological response to RSV infection that may lead to increased hospitalization with RSV at a younger age. Prior studies have also found the A allele associated with clinical presentation of allergic disease, including increased rates of asthma, atopic dermatitis, and IgE-mediated food allergy for both the AG and AA genotypes [52], but with a lower incidence of COVID19 [53]. Lastly, this variant has been associated with varying response to pegylated interferon-alpha and ribavirin treatment [54,55] and spontaneous clearance of hepatitis C infection [56,57], suggesting the influence on viral infection is not exclusive to respiratory diseases.

**Figure 2.**
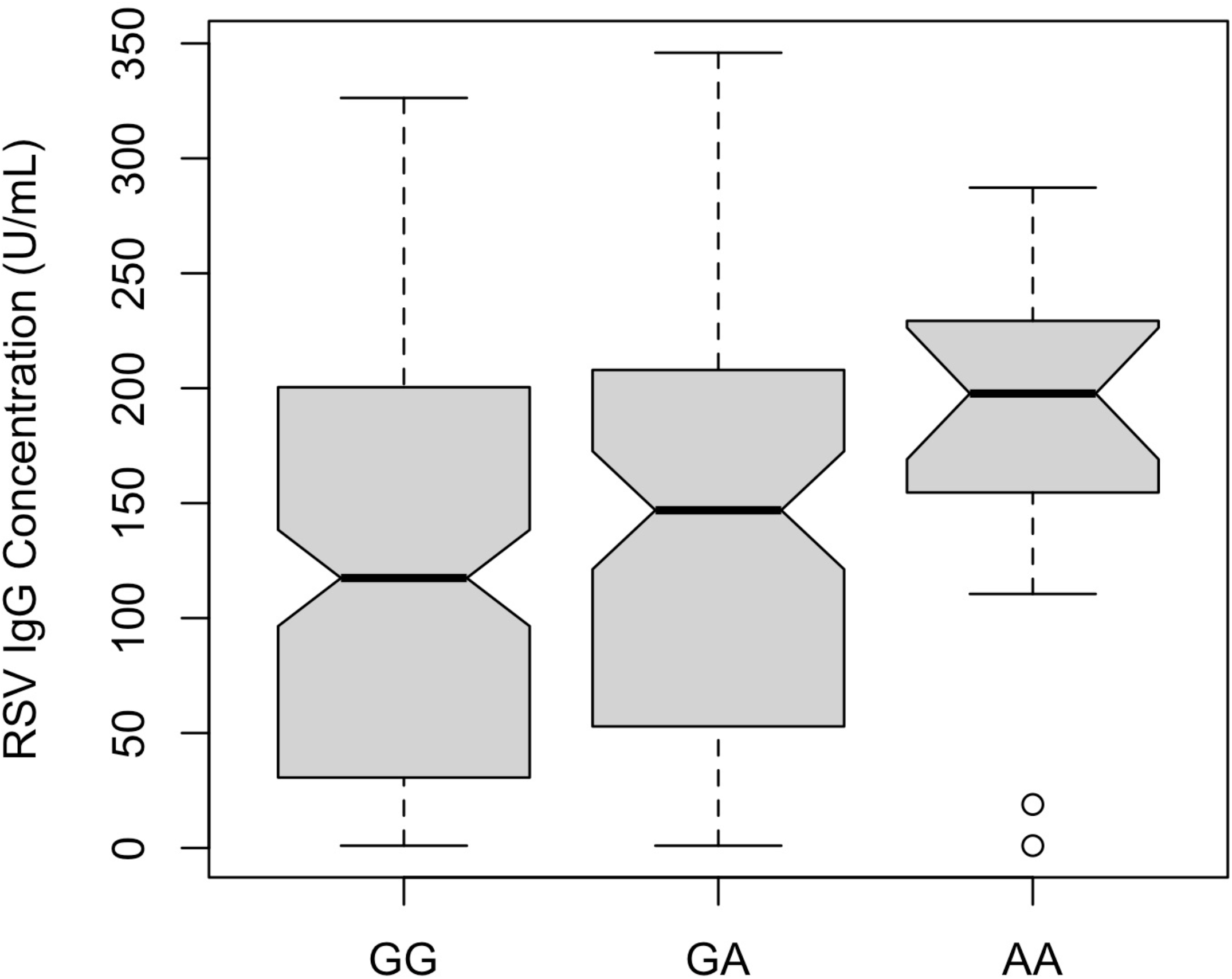
Median levels of RSV IgG concentration by genotype at rs12979860, an intronic variant in *IFNL4* that is associated with RSV IgG concentration in individuals with and without asthma (Table 6, p=0.0043). Individuals with two copies of the minor A allele (AA genotype) have significantly higher levels of RSV IgG as compared to individuals with two copies of the major G allele (GG genotype) (t-test, p=0.01).

We identified elevated total IgE concentrations (geometric mean 111.1 IU/ml) in both asthma cases and controls overall, similar to that reported in Alaskan Native children (geometric mean 122.1 kU/L) and in comparison to the Tucson Childhood Respiratory Study (geometric mean 36.3 kU/L) [58]. A Korean cohort of children between 12-13 years of age found a geometric mean total IgE of 90.6 kU/L and a 95 percentile of 991 kU/L [59]. High levels of IgE are consistent with a predominant inflammatory asthma phenotype in our study, and similar to that reported by Redding et al. in Alaskan Native children [58]. Elevated levels of total IgE suggest tendencies toward immune hyperreactivity and asthma at the community level, in accordance with reports of increased asthma prevalence in various other Indigenous communities [60,61]. Consistent with there being a tradeoff in IgE vs. IgG production, and preferential IgE production in Th2 cytokine environment [62,63], we indeed find lower levels of total IgE in individuals with high RSV-specific IgG concentrations as compared to individuals with low RSV concentrations (Table 2). However, total IgE was not predictive of RSV IgG in our multivariable model that included asthma status (Table 4), and may indicate a confounding correlation between asthma, % eosinophils, and serum total IgE on immunological response to RSV that requires further study.

We find children with high RSV IgG concentration tended to live in larger households, and had higher levels of cotinine levels indicating increased exposure to second-hand tobacco smoke. These observations are consistent with prior studies [51] that identified household crowding as a risk factor for RSV [64,65], including one study among Alaskan Native children [66]. Prior studies have also found a relationship between cotinine levels and increased risk of bronchiolitis and other complications of RSV infection [67–69]. While we could not examine the effect of household size and cotinine levels on clinical outcomes of RSV infection directly, our observation of higher cotinine levels in children with high RSV IgG concentration is important given that tobacco use and exposure is a serious public health concern in this community [70].

## Conclusion

In this study we report high RSV-specific IgG concentrations among a case-control study of asthma in American Indian children living in the Great Plains geographical area of the U.S. RSV-specific IgG concentrations were lower in children with asthma as compared to children without asthma during the winter months when RSV was likely circulating at higher levels in the community. Factors contributing to this observation include both genetic and environmental risk factors that require additional study.

## Data Availability

The data underlying the results of this study are owned and controlled by the Tribal entity that approved its collection. This fact is clearly stated in the Tribal resolution authorizing the research; and it must be recognized that this Tribal community is an independent, sovereign government, in control over research activities within their borders. Access to data and materials is accomplished by application to Dr. Lyle Best, who will arrange for further consultation with the appropriate Tribal partners. Approximately 2 to 3 months may be required. The authors received special access privileges to the data due to their relationship with the Tribal Government, however, interested researchers who apply for data access will be able to access the same data as the authors.

## Acknowledgements

We thank the study participants, Indian Health Service facilities, the Collaborative Research Center for American Indian Health, the Colorado Anschutz Research Genetics Organization (CARGO lab), and participating tribal community for their extraordinary cooperation and involvement, which has been critical to the success of this investigation. The views expressed in this paper are those of the authors and do not necessarily reflect those of the Indian Health Service. Funding was provided by the National Institute of Minority Health and Health Disparities, grant U54 MD008164.

## Supporting Information Captions

**S1 Fig.**
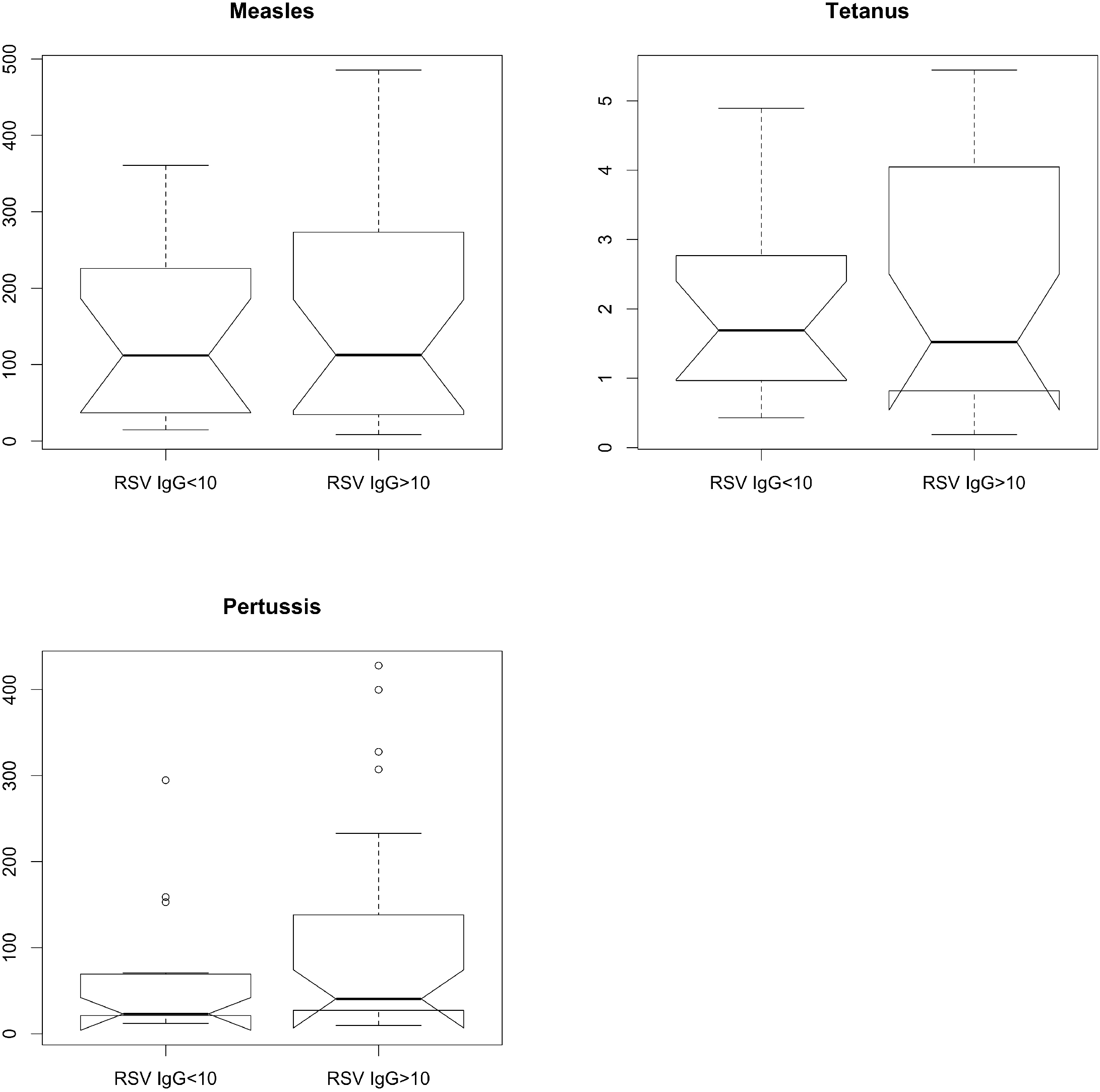
Boxplot showing the distribution of Measles, Pertussis, and Tetanus titers stratified by high (IgG>10 IU/mL, N=27) vs. low RSV IgG (IgG<10 IU/mL, N=16).

